# Sugar intake is associated with increased odds of depression and anxiety: Evidence from a cross-sectional study

**DOI:** 10.1101/2025.09.25.25336623

**Authors:** Christle Coxon, Mira Rufeger, Grace Hollamby, Deborah N Ashtree, Rebecca Orr, Melissa M Lane, Piril Hepsomali

## Abstract

**Background and Aims:** The current study aimed to assess the associations between (i) total and specific sugar intake, (ii) dietary exposures that met Global Burden of Diseases, Injuries, and Risk Factors Study (GBD) selection criteria for risk factors, and depression and anxiety.

**Methods:** In this online cross-sectional study that took place between 2022-2024 in the United Kingdom, 377 participants between the ages of 18-66 years (M=26.09; SD=8.48) completed the: EPIC-Norfolk Food Frequency Questionnaire from which overall sugar intake, specific sugar intake (fructose, galactose, glucose, lactose, maltose, and sucrose) and GBD dietary exposures were derived; and the Depression, Anxiety and Stress Scales from which likely depression and anxiety cases were identified.

**Results:** The prevalence of depression and anxiety was 12.5% and 16.4%, respectively. Separate logistic regression models assessing the associations between dietary intake and depression) and anxiety revealed that total sugar intake was associated with greater odds of depression (OR: 1.01, 95%CI 1.00 to 1.02) and anxiety (OR: 1.01, 95%CI 1.01 to 1.02). Specifically, higher sucrose intake was associated with greater odds of anxiety (OR: 1.02, 95%CI 1.00 to 1.05), while higher intake of sugar-sweetened beverages was associated with increased odds of both depression (OR: 1.00, 95%CI 1.00 to 1.01) and anxiety (OR: 1.00, 95%CI 1.00 to 1.01).

**Conclusion:** While higher overall sugar intake was associated with both depression and anxiety, sucrose intake emerged as a specific factor associated with increased odds of anxiety, and higher sugar-sweetened beverages intake with depression and anxiety, warranting further investigation into their potential role in mental health outcomes.

## 1. Introduction

Common mental disorders, such as depressive- and anxiety-related disorders are highly comorbid (1) and globally prevalent, affecting more than 300 million and 250 million people, respectively (2). The associated social, economic, and health burden of depression and anxiety costs the world economy trillions of dollars each year (3). Thus, preventive and treatment strategies are needed to tackle this major public health problem and ultimately improve the lives of those affected. However, pharmacological treatments for depression and anxiety has overestimated efficacy (4). Given these challenges, exploring alternative targets for depression and anxiety prevention and treatment has become increasingly imperative.

Diet is a leading risk factor for non-communicable diseases, as acknowledged by the Global Burden of Diseases, Injuries, and Risk Factors Study (GBD) (5), and may be protective against depression and anxiety outcomes (6, 7). This may occur via modulating numerous physiological channels, including regulation of systemic inflammation, and promotion of short chain fatty acids and neurotransmitters (gamma-aminobutyric acid (GABA), serotonin, dopamine) (8, 9). Therefore, dietary interventions could provide an effective strategy to improve and prevent depressive and anxiety-related outcomes (10, 11).

Furthermore, adhering to healthful dietary patterns may potentially reduce the risks of developing depression and anxiety. For example, diets characterised by high intakes of vegetables, fruits, nuts, legumes, and unprocessed cereals, and low intakes of meat and dairy products are associated with lower odds/risk of depression and anxiety (12). Further, diets higher in saturated fats, sodium, and sugar have been linked to higher odds for depression and anxiety (13). Recent meta-analyses have further confirmed the link between depression and anxiety and Western-type foods (14), ultra-processed foods (15), and sugar-sweetened beverages (16). One potential nutrient/food group these dietary patterns have in common is high sugar intake.

Globally, sugar and sugar-sweetened beverage intake is increasing (17). There is now compelling evidence to suggest a causal relationship between sugar-sweetened beverage intake and many health outcomes, including high body mass index, ischemic heart disease and type 2 diabetes (17). Currently, the role of sugar-sweetened beverage intake in depression and anxiety has not been well studied. Still, a meta-analysis of observational studies found that drinking the equivalent of two cups of cola per day increased the risk of depression by up to five percent (16). However, in a recent umbrella review of many meta-analyses (18), the meta-analysis by Hu et al., 2019 (16) was classified as low quality as per the Grading of Recommendations, Assessment, Development and Evaluation criteria due to significant risk of bias. Furthermore, accordingly, total dietary sugar intake, but not specific sugar-sweetened beverages, has been cross-sectionally linked to higher odds of depression (19). Supporting this, a recent study by Graybeal et al. (20) identified added and relative sugar as the only dietary factors associated with depression, with associations also linked to mental health medication use and mediated by emotional eating and craving control.

Together, these findings highlight the need for further rigorous studies to explore dietary intake, particularly sugar intake, in relation to depression and anxiety.

It is important to note that the relationship between sugar intake (and excess intake of other nutrients, such as processed meats) and mental health symptoms may be bidirectional, such that individuals experiencing depressive or anxious symptoms may be more likely to engage in deregulatory/coping behaviours such as higher emotional eating and lower craving (20, 21), leading to higher consumption of sugar and other unhealthy food items (22). Furthermore, factors such as overall health status and body mass index (BMI) likely modulate this relationship, influencing both dietary habits and mental health outcomes (23). Although aforementioned evidence clearly highlights the contributions of dietary exposures to depression and anxiety, particularly sugar and sugar-sweetened beverage intake, the GBD does not currently generate risk-outcome estimates for diet and depression and anxiety. To address this gap, we aimed to assess the associations of (i) total and specific sugar intake, (ii) sugar-sweetened beverage intake, as defined by the GBD criteria for risk factors, and (iii) other dietary exposures that meet GBD selection criteria for risk factors with depression and anxiety.

## 2. Methods

### 2.1 Study Design

This study was conducted according to the guidelines laid down in the Declaration of Helsinki and all procedures involving research study participants were approved by the Ethical committee at the University of Roehampton (Reference: PSYC/22420). A convenience sample of 532 participants (mainly university students from the University of Roehampton, United Kingdom) were recruited through University web portals, social media, and word of mouth between 2022-2024. All participants provided a written informed consent to participate in the research. They completed a web-based survey on Qualtrics (Qualtrics, Provo, UT). The exclusion criteria were: any history of, or taking medication for, psychiatric disorders or diseases (ADHD, depression, anxiety, or mood disorders), sleep disorders, or neurological disorders or diseases (stroke, head injury, epilepsy, seizures, brain tumours, brain surgery, Parkinson’s Disease). After data cleaning (see Note S1 in Supplementary Materials), a final sample of 377 participants (306 females, 65 males, 6 preferred not to say) between the ages of 18 - 66 years (M = 26.09; SD = 8.48) were included in the analysis. This study was reported in line with the Strengthening the Reporting of Observational Studies in Epidemiology (STROBE) statement and checklist for observational studies (24) (see the Supplementary STROBE checklist).

### 2.2 Exposure Variables

In order to define dietary intake according to the GBD study, we followed the Global burden of disease Lifestyle And mental Disorder (GLAD) Project guidelines (25). Specifically, we used various dietary exposure variables (intakes in grams per day [g/d] of: fruit; vegetables; legumes; whole grains; nuts and seeds; milk; red meat; processed meat; sugar-sweetened beverages; fibre; calcium; sodium; as well as percentage of total daily energy intake of polyunsaturated fatty acid [PUFA] and trans-fat). Additionally, we included intakes (g/d) of total dietary sugar, as well as specific sugars: fructose, galactose, glucose, lactose, maltose, and sucrose. Dietary data were collected using the European Prospective Investigation into Cancer (EPIC) Norfolk Food Frequency Questionnaire (FFQ) (26) and analysed using the FETA Software (27), reflecting habitual intake of the nutrients of interest. Trans-fat intake was estimated by linking FFQ data with a nutrient database specific to the EPIC Norfolk study (26) (see Note S2 in Supplementary Materials).

### 2.3 Outcome Variables

The scores on the depression and anxiety subscales of the Depression, Anxiety, and Stress Scales (DASS) (28) were used to identify individuals likely with and without depression and anxiety, respectively. The DASS consists of 42 questions, such as “I felt terrified”, requiring participants to note how frequently they identified with this statement over the past week. The DASS is scored on a four-point Likert scale, from zero (“did not apply to me at all”) to three (“applied to me very much, or most of the time”). Depression and anxiety were dichotomised using validated cut-offs for the relevant subscales: Participants with a score of ≥21 on the depression scale were identified as likely having depression (28), and participants with a score of ≥15 on the anxiety scale were identified as likely having anxiety (28).

### 2.4 Statistical Analysis

All analyses were conducted using IBM SPSS 29.0.1.0. Depression and anxiety present and absent groups were compared on demographic and dietary measures by using Mann-Whitney-U (Benjamini-Hochberg-corrected), chi-square (Bonferroni-corrected) or independent sample t-tests (Benjamini-Hochberg-corrected). We fitted logistic regression models following the GLAD project protocol (25) to assess the association of (i) sugar-sweetened beverage intake (primary analysis), (ii) total and specific sugar intake, and (iii) dietary intake, as defined above, and odds of depression and anxiety. All dietary variables were modelled as continuous variables, with the outcomes modelled as binary (depression- and anxiety-present or absent). Odds ratios and their corresponding 95% confidence intervals were estimated. We used three levels of adjustment - model 1: unadjusted; model 2: adjusted for age, sex, income; and model 3: as in model 2, with BMI (kg/m^2^). Sensitivity analyses were performed using (i) energy-adjusted versions of all dietary variables (29) and (ii) the whole sample, before exclusion in order to maximise power and improve generalisability. All significance tests were two-tailed using p < 0.05 as the level of significance and corrected by using the Benjamini-Hochberg method.

## 3. Results

### 3.1 Sample Characteristics

Participant characteristics are reported in Table 1. Of 377 participants, 12.5% (n=47) and 16.4% (n=62) had depression and anxiety, respectively. Groups did not differ in age, BMI, or energy intake. Participants who identified their gender as ‘other/prefer not to answer’ were more likely to have both depression and anxiety. Individuals who earned less than £18,000 per year were more likely to have depression.

**Table 1.** Participant characteristics across depression absent, depression present, anxiety absent, and anxiety present groups.

|  | Depression |  |  |  | Anxiety |  |  |  |
| --- | --- | --- | --- | --- | --- | --- | --- | --- |
|  | Absent (n=330) | Present (n=47) |  |  | Absent (n=315) | Present (n=62) |  |  |
|  | Median (IQR) | Median (IQR) | M-W U | Padj | Median (IQR) | Median (IQR) | M-W U | Pad |
| Age (years) | 23.0 (20.0-32.0) | 21.0 (20.0-24.0) | 5703.0 | 0.08 | 23.0 (20.0-32.3) | 20.0 (19.0-23.8) | 6309.0 | 0.08 |
| Body Mass Index (kg/m<sup>2</sup>) | 23.17 (20.76-26.21) | 23.8 (19.76-25.91) | 6360.5 | 0.26 | 23.17 (20.75-25.97) | 23.04 (19.79-27.50) | 8475.0 | 0.26 |
| Energy (kcal/day) | 1466.46 (1089.96-1882.31) | 1512.40 (1104.63-2141.61) | 8761.0 | 0.17 | 1442.10 (1083.26-1822.09) | 1546.83 (1160.28-2371.23) | 11647.0 | 0.17 |
|  | N (%) | N (%) | X<sup>2</sup> | Padj | N (%) | N (%) | X<sup>2</sup> | Padj |
| Gender |  |  | 16.82 | <0.001 |  |  | 23.23 | <0.001 |
| Male | 59a (17.9) | 6a (12.8) |  |  | 60a (19.0) | 5b (8.1) |  |  |
| Female | 269a (81.5) | 37a (78.7) |  |  | 254a (80.6) | 52a (83.9) |  |  |
| Other/prefer not to answer | 2a (0.6) | 4b (8.5) |  |  | 1a (0.3) | 5b (8.1) |  |  |
| Income |  |  | 14.12 | 0.02 |  |  | 8.53 | 0.13 |
| Less than £18,000 | 50a (15.2) | 15b (31.9) |  |  | 52a (16.5) | 13a (21.0) |  |  |
| £18,000 to £30,999 | 54a (16.4) | 7a (14.9) |  |  | 47a (14.9) | 14a (22.6) |  |  |
| £31,000 to £51,999 | 68a (20.6) | 8a (17.0) |  |  | 68a (21.6) | 8a (12.9) |  |  |
| £52,000 to £100,000 | 65a (19.7) | 4a (8.5) |  |  | 63a (20.0) | 6a (9.7) |  |  |
| More than £100,000 | 24a (7.3) | 0a (0.0) |  |  | 20a (6.3) | 4a (6.5) |  |  |
| Other* | 69a (20.9) | 13a (27.7) |  |  | 65a (20.6) | 17a (27.4) |  |  |

IQR: Interquartile range; M-W: Mann Whitney; \*Do not know/prefer not to answer; Each subscript letter denotes a subset of Depression and Anxiety categories whose column proportions do not differ significantly from each other at the .05 level.

Average dietary intakes are presented in Table 2, separately for participants with and without depression or anxiety. Total sugar, as well intakes of sugar-sweetened beverages were higher in participants with depression and anxiety. Red meat intake was higher in the depression-present group. Participants with anxiety had higher sodium, maltose, and sucrose intakes compared to participants without anxiety.

**Table 2.** Dietary intakes across depression absent/present and anxiety absent/present groups.

|  | Depression |  |  |  |  |  | Anxiety |  |  |  |  |  |
| --- | --- | --- | --- | --- | --- | --- | --- | --- | --- | --- | --- | --- |
|  | Absent (n=330) |  | Present (n=47) |  | *t* | *Padj* | Absent (n=315) |  | Present (n=62) |  | *t* | *Padj* |
|  | *Mean* | *SD* | *Mean* | *SD* |  |  | *Mean* | *SD* | *Mean* | *SD* |  |  |
| Fruits (g/day) | 174.82 | 169.70 | 200.18 | 212.31 | -0.93 | 0.14 | 171.64 | 163.34 | 210.20 | 226.15 | -1.59 | 0.11 |
| Vegetables (g/day) | 240.72 | 178.67 | 230.55 | 169.83 | 0.37 | 0.23 | 238.53 | 177.83 | 244.15 | 176.60 | -0.23 | 0.23 |
| Legumes (g/day) | 18.65 | 21.56 | 15.18 | 12.91 | 1.075 | 0.13 | 18.29 | 21.56 | 17.84 | 15.73 | 0.16 | 0.26 |
| Wholegrains (g/day) | 230.00 | 151.65 | 240.79 | 141.92 | -0.46 | 0.20 | 225.77 | 148.88 | 259.66 | 155.68 | -1.63 | 0.13 |
| Nuts & Seeds (g/day) | 8.65 | 13.04 | 7.68 | 10.32 | 0.49 | 0.18 | 8.81 | 13.28 | 7.12 | 9.39 | 0.96 | 0.21 |
| Milk (g/day) | 254.42 | 180.25 | 261.66 | 192.48 | -0.26 | 0.25 | 248.51 | 174.62 | 289.90 | 211.62 | -1.65 | 0.09 |
| Red meat (g/day) | 36.58 | 42.69 | 55.88 | 59.27 | -2.75 | **0.04** | 37.69 | 43.81 | 45.60 | 52.89 | -1.26 | 0.16 |
| Processed meat (g/day) | 76.23 | 16.58 | 83.50 | 25.33 | -2.61 | 0.05 | 76.69 | 16.95 | 79.43 | 22.74 | -1.10 | 0.20 |
| Sugar-sweetened beverages (g/day) | 39.11 | 59.29 | 81.03 | 113.26 | -3.94 | **0.02** | 39.67 | 60.47 | 68.06 | 101.56 | -2.97 | **0.02** |
| Fibre (g/day) | 14.11 | 7.24 | 14.58 | 7.83 | -0.41 | 0.21 | 13.95 | 7.00 | 15.27 | 8.65 | -1.30 | 0.14 |
| Calcium (g/day) | 0.71 | 0.30 | 0.74 | 0.32 | -0.62 | 0.16 | 0.70 | 0.29 | 0.79 | 0.34 | -2.28 | 0.05 |
| Sodium (g/day) | 2.07 | 0.91 | 2.37 | 1.27 | -1.96 | 0.07 | 2.05 | 0.89 | 2.40 | 1.24 | -2.58 | **0.04** |
| PUFA (%*) | 6.43 | 1.98 | 6.03 | 1.48 | 1.34 | 0.09 | 6.43 | 1.99 | 6.12 | 1.54 | 1.16 | 0.18 |
| Trans-fats (%*) | 0.13 | 0.05 | 0.14 | 0.07 | -1.12 | 0.11 | 0.13 | 0.05 | 0.15 | 0.07 | -1.96 | 0.07 |
| Sugars total (g/day) | 82.54 | 39.66 | 104.76 | 62.99 | -3.30 | **0.04** | 81.11 | 38.73 | 106.63 | 59.44 | -4.29 | **0.04** |
| *Fructose (g/day)* | 16.11 | 9.60 | 19.48 | 11.40 | -2.20 | 0.14 | 15.87 | 9.23 | 19.84 | 12.30 | -2.91 | 0.11 |
| *Galactose (g/day)* | 0.44 | 0.72 | 0.29 | 0.44 | 1.34 | 0.18 | 0.43 | 0.71 | 0.37 | 0.55 | 0.67 | 0.18 |
| *Glucose (g/day)* | 15.59 | 8.69 | 19.61 | 11.33 | -2.85 | 0.11 | 15.44 | 8.47 | 19.40 | 11.52 | -3.16 | 0.07 |
| *Lactose (g/day)* | 9.95 | 6.91 | 10.99 | 8.02 | -0.95 | 0.21 | 9.73 | 6.63 | 11.85 | 8.72 | -2.18 | 0.14 |
| *Maltose (g/day)* | 2.19 | 1.65 | 3.22 | 3.30 | -3.44 | 0.07 | 2.17 | 1.66 | 3.07 | 2.96 | -3.37 | **0.04** |
| *Sucrose (g/day)* | 36.06 | 20.50 | 48.77 | 38.09 | -3.49 | 0.07 | 35.39 | 20.25 | 49.12 | 34.61 | -4.26 | **0.04** |

\* percentage of total daily energy intake

### 3.2. Associations between sugar intake and depression and anxiety

Results are presented in Table 3. Higher total sugar intake was associated with greater odds of depression and anxiety in the unadjusted (Model 1) but not in the adjusted models (Model 2 and Model 3). Higher galactose intake was associated with greater odds of depression in the unadjusted model (Model 1), but not in the adjusted models (Model 2 and Model 3). Higher sucrose intake was associated with greater odds of anxiety in the unadjusted model (Model 1), but not in the adjusted models (Model 2 and Model 3).

**Table 3.** Logistic regression analyses of the associations of sugar intake with depression and anxiety.

| Depression |  |  |  |  |  |  |  |  |  |  |  |  |
| --- | --- | --- | --- | --- | --- | --- | --- | --- | --- | --- | --- | --- |
|  | Model 1*** |  |  |  | Model 2*** |  |  |  | Model 3*** |  |  |  |
|  | OR | p | 95%CI<br>Lower/Upper |  | OR | P<sub>adj</sub> | 95%CI<br>Lower/Upper |  | OR | P<sub>adj</sub> | 95%CI<br>Lower/Upper |  |
| Sugars total | 1.01** | 0.008 | 1.00 | 1.02 | 1.01 | 0.07 | 1.00 | 1.02 | 1.01* | 0.04 | 1.00 | 1.02 |
|  | Model 1*** |  |  |  | Model 2*** |  |  |  | Model 3*** |  |  |  |
|  | OR | P<sub>adj</sub> | 95%CI<br>Lower/Upper |  | OR | P<sub>adj</sub> | 95%CI<br>Lower/Upper |  | OR | P<sub>adj</sub> | 95%CI<br>Lower/Upper |  |
| Fructose | 0.97 | 0.17 | 0.83 | 1.13 | 0.97 | 0.19 | 0.82 | 1.14 | 0.96 | 0.18 | 0.81 | 1.14 |
| Galactose | 0.53 | 0.04 | 0.26 | 1.08 | 0.58 | 0.11 | 0.28 | 1.19 | 0.57 | 0.10 | 0.27 | 1.19 |
| Glucose | 1.05 | 0.13 | 0.87 | 1.27 | 1.07 | 0.17 | 0.88 | 1.31 | 1.07 | 0.16 | 0.87 | 1.32 |
| Lactose | 1.01 | 0.21 | 0.96 | 1.06 | 1.00 | 0.20 | 0.96 | 1.06 | 1.00 | 0.24 | 0.95 | 1.06 |
| Maltose | 0.99 | 0.25 | 0.69 | 1.41 | 0.96 | 0.23 | 0.66 | 1.41 | 0.96 | 0.21 | 0.65 | 1.41 |
| Sucrose | 1.01 | 0.08 | 0.99 | 1.04 | 1.01 | 0.16 | 0.98 | 1.04 | 1.01 | 0.15 | 0.98 | 1.04 |
| Anxiety |  |  |  |  |  |  |  |  |  |  |  |  |
|  | Model 1*** |  |  |  | Model 2*** |  |  |  | Model 3*** |  |  |  |
|  | OR | p | 95%CI<br>Lower/Upper |  | OR | P<sub>adj</sub> | 95%CI<br>Lower/Upper |  | OR | P<sub>adj</sub> | 95%CI<br>Lower/Upper |  |
| Sugars total | 1.01 | <0.001 | 1.01 | 1.02 | 1.01 | 0.022 | 1.00 | 1.02 | 1.01 | 0.020 | 1.01 | 1.02 |
|  | Model 1*** |  |  |  | Model 2*** |  |  |  | Model 3*** |  |  |  |
|  | OR | P<sub>adj</sub> | 95%CI<br>Lower/Upper |  | OR | P<sub>adj</sub> | 95%CI<br>Lower/Upper |  | OR | P<sub>adj</sub> | 95%CI<br>Lower/Upper |  |
| Fructose | 1.10 | 0.08 | 0.97 | 1.26 | 1.11 | 0.09 | 0.96 | 1.30 | 1.13 | 0.10 | 0.97 | 1.31 |
| Galactose | 0.76 | 0.17 | 0.44 | 1.30 | 0.76 | 0.19 | 0.41 | 1.41 | 0.77 | 0.19 | 0.42 | 1.43 |
| Glucose | 0.89 | 0.13 | 0.75 | 1.05 | 0.88 | 0.11 | 0.73 | 1.07 | 0.87 | 0.12 | 0.72 | 1.06 |
| Lactose | 1.01 | 0.25 | 0.97 | 1.06 | 1.02 | 0.20 | 0.97 | 1.08 | 1.02 | 0.21 | 0.97 | 1.08 |
| Maltose | 1.11 | 0.21 | 0.80 | 1.54 | 1.04 | 0.23 | 0.72 | 1.51 | 1.04 | 0.24 | 0.72 | 1.50 |
| Sucrose | 1.02 | 0.04 | 1.00 | 1.05 | 1.03 | 0.06 | 1.00 | 1.05 | 1.03 | 0.06 | 1.00 | 1.05 |

\* p≤ 0.05, \*\* p≤ 0.01, p≤ 0.001
Model 1: unadjusted; Model 2: adjusted for age, sex, income; and Model 3: as in model 2, with body mass index (BMI; kg/m<sup>2</sup>)

Results from sensitivity analyses are mostly consistent with results reported here, apart from the lack of galactose and depression association (see Supplementary Table 3 in Supplementary Materials). Sensitivity analyses of the associations of sugar intake with depression and anxiety in the whole sample (prior to participant exclusion) are nearly identical with results reported here, however, in these models, higher total sugar intake was associated with greater odds of depression and anxiety in both of the adjusted models (see Supplementary Table 4 in Supplementary Materials).

### 3.3. Associations between sugar-sweetened beverages and depression and anxiety

Logistic regression analyses of the associations between dietary variables (intakes in grams per day [g/d] of: fruit; vegetables; legumes; whole grains; nuts and seeds; milk; red meat; processed meat; sugar-sweetened beverages; fibre; calcium; sodium; as well as percentage of total daily energy intake of PUFA and trans-fat) and depression and anxiety are reported in Table 4. In the unadjusted models (Model 1), dietary intake was not associated with depression or anxiety. However, in adjusted models, higher sugar-sweetened beverage intake was associated with greater odds of depression (Model 2 and Model 3) and anxiety (Model 2).

**Table 4.** Logistic regression analyses of the associations of dietary variables with depression and anxiety.

| Depression |  |  |  |  |  |  |  |  |  |  |  |  |
| --- | --- | --- | --- | --- | --- | --- | --- | --- | --- | --- | --- | --- |
|  | Model 1 |  |  |  | Model 2** |  |  |  | Model 3** |  |  |  |
|  | OR | *P*<sub>adj</sub> | 95%CI<br>Lower/Upper |  | OR | *P*<sub>adj</sub> | 95%CI<br>Lower/Upper |  | OR | *P*<sub>adj</sub> | 95%CI<br>Lower/Upper |  |
| Fruits | 1.00 | 0.14 | 1.00 | 1.00 | 1.00 | 0.21 | 1.00 | 1.00 | 1.00 | 0.21 | 1.00 | 1.00 |
| Vegetables | 1.00 | 0.21 | 1.00 | 1.00 | 1.00 | 0.22 | 1.00 | 1.00 | 1.00 | 0.22 | 1.00 | 1.00 |
| Legumes | 0.98 | 0.04 | 1.00 | 1.01 | 0.98 | 0.09 | 1.00 | 1.01 | 0.98 | 0.08 | 1.00 | 1.01 |
| Wholegrains | 1.00 | 0.20 | 1.00 | 1.00 | 1.00 | 0.12 | 1.00 | 1.00 | 1.00 | 0.11 | 1.00 | 1.00 |
| Nuts & Seeds | 1.02 | 0.09 | 0.98 | 1.05 | 1.03 | 0.07 | 0.99 | 1.06 | 1.03 | 0.07 | 0.99 | 1.06 |
| Milk | 1.00 | 0.23 | 1.00 | 1.00 | 1.00 | 0.16 | 1.00 | 1.00 | 1.00 | 0.15 | 1.00 | 1.00 |
| Red meat | 1.01 | 0.05 | 1.00 | 1.01 | 1.01 | 0.10 | 1.00 | 1.01 | 1.01 | 0.10 | 1.00 | 1.01 |
| Processed meat | 1.00 | 0.18 | 0.98 | 1.02 | 1.01 | 0.15 | 1.00 | 1.03 | 1.01 | 0.14 | 1.00 | 1.03 |
| Sugar-sweetened beverages | 1.01 | 0.02 | 1.00 | 1.01 | 1.00 | 0.04 | 1.00 | 1.01 | 1.00 | 0.04 | 1.00 | 1.01 |
| Fibre | 0.10 | 0.25 | 0.86 | 1.16 | 1.01 | 0.24 | 0.86 | 1.19 | 1.01 | 0.24 | 0.86 | 1.19 |
| Calcium | 0.59 | 0.13 | 0.09 | 3.98 | 0.75 | 0.19 | 0.09 | 6.13 | 0.75 | 0.18 | 0.09 | 6.12 |
| Sodium | 0.96 | 0.07 | 0.76 | 1.20 | 0.99 | 0.13 | 0.78 | 1.25 | 0.99 | 0.13 | 0.78 | 1.25 |
| PUFA | 1.37 | 0.16 | 0.72 | 2.61 | 1.34 | 0.25 | 0.66 | 2.71 | 1.34 | 0.25 | 0.66 | 2.72 |
| Trans-fats | 2.97 | 0.11 | 0.21 | 41.57 | 1.60 | 0.18 | 0.12 | 22.31 | 1.58 | 0.17 | 0.11 | 22.09 |
| Anxiety |  |  |  |  |  |  |  |  |  |  |  |  |
|  | Model 1 |  |  |  | Model 2*** |  |  |  | Model 3*** |  |  |  |
|  | OR | *P*<sub>adj</sub> | 95%CI<br>Lower/Upper |  | OR | *P*<sub>adj</sub> | 95%CI<br>Lower/Upper |  | OR | *P*<sub>adj</sub> | 95%CI<br>Lower/Upper |  |
| Fruits | 1.00 | 0.09 | 1.00 | 1.00 | 1.00 | 0.24 | 1.00 | 1.00 | 1.00 | 0.21 | 1.00 | 1.00 |
| Vegetables | 1.00 | 0.25 | 1.00 | 1.00 | 1.00 | 0.21 | 1.00 | 1.00 | 1.00 | 0.24 | 1.00 | 1.00 |
| Legumes | 0.99 | 0.07 | 1.00 | 1.01 | 0.99 | 0.07 | 1.00 | 1.01 | 0.99 | 0.07 | 0.97 | 1.01 |
| Wholegrains | 1.00 | 0.11 | 1.00 | 1.00 | 1.00 | 0.22 | 1.00 | 1.00 | 1.00 | 0.25 | 1.00 | 1.00 |
| Nuts & Seeds | 1.00 | 0.14 | 1.00 | 1.03 | 1.01 | 0.10 | 1.00 | 1.05 | 1.01 | 0.08 | 1.00 | 1.05 |
| Milk | 1.00 | 0.21 | 1.00 | 1.00 | 1.00 | 0.13 | 1.00 | 1.00 | 1.00 | 0.11 | 1.00 | 1.00 |
| Red meat | 1.00 | 0.23 | 0.99 | 1.01 | 1.00 | 0.15 | 0.99 | 1.01 | 1.00 | 0.15 | 0.99 | 1.01 |
| Processed meat | 1.00 | 0.20 | 1.00 | 1.02 | 1.00 | 0.25 | 0.98 | 1.02 | 1.00 | 0.22 | 1.00 | 1.02 |
| **Sugar-sweetened beverages** | 1.00 | 0.02 | 1.00 | 1.01 | **1.00** | 0.04 | 1.00 | 1.01 | 1.00 | 0.06 | 1.00 | 1.01 |
| **Fibre** | 1.01 | 0.18 | 0.89 | 1.14 | 1.02 | 0.19 | 0.89 | 1.16 | 1.01 | 0.19 | 0.88 | 1.16 |
| **Calcium** | 1.13 | 0.16 | 0.22 | 5.81 | 1.59 | 0.12 | 0.25 | 9.98 | 1.67 | 0.13 | 0.27 | 10.33 |
| **Sodium** | 0.96 | 0.05 | 0.79 | 1.18 | 1.03 | 0.16 | 0.83 | 1.27 | 1.02 | 0.17 | 0.83 | 1.25 |
| **PUFA** | 1.22 | 0.13 | 0.70 | 2.12 | 1.11 | 0.18 | 0.59 | 2.09 | 1.14 | 0.18 | 0.60 | 2.14 |
| **Trans-fats** | 4.09 | 0.04 | 0.35 | 48.50 | 2.23 | 0.09 | 0.18 | 27.55 | 2.02 | 0.14 | 0.16 | 25.47 |

\* $p \leq 0.05$ , \*\* $p \leq 0.01$ , $p \leq 0.001$
*Model 1: unadjusted; Model 2: adjusted for age, sex, income; and Model 3: as in model 2, with body mass index (BMI; kg/m<sup>2</sup>)*

Sensitivity analyses regarding depression produced an identical pattern of results to those reported here, but sugar-sweetened beverage intake was not associated with anxiety in any of the models (see Supplementary Table 5 in Supplementary Materials). Sensitivity analyses of the associations of sugar intake with depression and anxiety in the whole sample (prior to participant exclusion) revealed that higher sugar-sweetened beverage was associated with greater odds of depression and anxiety in the unadjusted models (Models 1). In both adjusted models (Model 2 and Model 3), higher sugar-sweetened beverage intake was associated with greater odds of depression (see Supplementary Table 6 in Supplementary Materials).

### 3.4. Associations between dietary intake and depression, and anxiety

Other GBD-defined dietary exposures (intakes in grams per day [g/d] of: fruit; vegetables; legumes; whole grains; nuts and seeds; milk; red meat; processed meat; sugar-sweetened beverages; fibre; calcium; sodium; as well as percentage of total daily energy intake of PUFA and trans-fat) were not associated with depression or anxiety (Table 4). Results were consistent in sensitivity analyses (see Supplementary Table 5 in Supplementary Materials). Sensitivity analyses in the whole sample (prior to participant exclusion) revealed that higher red meat intakes were associated with greater odds of depression and anxiety in the unadjusted models (Models 1). In all models, calcium intake was associated with greater odds of anxiety. (see Supplementary Table 6 in Supplementary Materials).

## 4. Discussion

The current cross-sectional study revealed distinct patterns of dietary intakes in individuals with depression or anxiety compared to participants without depression or anxiety. Participants with depression or anxiety had greater intakes of sugar-sweetened beverages, total dietary sugar, fructose, glucose, maltose, and sucrose compared to those without depression and anxiety. Higher sugar-sweetened beverage intake was associated with increased odds of depression. Furthermore, higher dietary sugar intake was associated with increased odds of depression and anxiety. However, only sucrose intake was associated with increased odds for anxiety. It is important to emphasise that the observed associations, although statistically significant, are characterised by odds ratios very close to 1.00 and narrow confidence intervals, indicating marginal effects with limited clinical relevance. We recognise that such small effect sizes, despite reaching statistical significance, may have minimal practical or clinical implications.

Our results are concordant with previous research demonstrating associations between sugar-sweetened beverage intake and depression (16), possibly due to the link between dietary sugar intake and depression (18, 19). There are several plausible biological explanations for these associations. Sugar consumption might cause depression through hyperfunction of the hypothalamic-pituitary-adrenal (HPA) axis, leading to excessive cortisol release and altered feedback inhibition mediated by the glucocorticoid receptor (30). This is supported by observations from pre-clinical studies showing that reported fructose intake in peri-adolescence can stimulate the HPA axis and induce increased depressive-like behaviour in adult rats (31). Additional studies have shown that rats who were fed high-fat high-sugar diets exhibited a decrease in the brain-derived neurotrophic factor (BDNF) (32). Human studies have subsequently shown that low serum and plasma BDNF levels are associated with depression and anxiety (33).

Another biological mechanism linking sugar intake with depression is via gut-brain interactions (8). Previous studies have demonstrated that the gut microbiota likely influence the development of mental disorders, including depression (34). Specifically, intake of sugar-sweetened beverages and dietary sugar intake could lead to depression or anxiety by disrupting gut microbiota composition and function (35), possibly through increased abundances of lactic acid-producing bacteria resulting in lactate accumulation, lower levels of the butyrate-producing bacteria, and/or greater utilisation of glutamate (i.e. depletion) and increased synthesis of GABA (36).

Because sugar-sweetened beverages and dietary sugar both have a high glycaemic index, high consumption may potentially lead to insulin resistance (37). Insulin resistance could influence the likelihood of developing depression and anxiety via altered dopamine turnover (38) or via shared pro-inflammatory pathways (39). Increased low-grade inflammation induced by sugar-sweetened beverages and/or dietary sugar intake (40) could subsequently increase risk of depression and anxiety, possibly via altering production, metabolism, and transport of dopamine, glutamate, and serotonin that synergistically affect mood (41). These shared pathways between mental and physical health mean sugar intake can directly influence whether someone develops depression and anxiety but may also act secondarily through other physical health conditions. Specifically, higher body mass index and diabetes caused by high sugar intake (42) could subsequently increase the likelihood of depression and anxiety (17).

Finally, it is important to note that the abovementioned biological mechanisms are mostly related to the sugar content of sugar-sweetened beverages, however, other ingredients, such as caffeine and non-nutritive sweeteners/sugar substitutes (aspartame, saccharine, sucralose, neotame, acesulfame-K, and stevia), have also been shown to be linked to higher levels of depression and anxiety directly or indirectly (through increasing the risk of non-communicable diseases) (43–45).

Possible psychosocial explanations for these associations should not be overlooked. Individuals who consume higher amounts of sugar-sweetened beverages and/or dietary sugar are more likely to have a higher BMI (42). The adverse effects of having a high BMI on psychological wellbeing may be partly explained by weight discrimination (46). Additionally, consumption of higher amounts of sugar-sweetened beverages and/or dietary sugar is associated with a variety of socio-economic factors (such as low socio-economic status) (47) that are known to be risk factors for developing depression and/or anxiety (48). However, the current study partly accounted for this by adjusting for income, suggesting that other factors may explain the observed associations. Finally, individuals with depression and anxiety often exhibit poorer dietary quality and unhealthy eating behaviours, including emotional eating (21), and so they may be more likely to select sweet foods/sugar-sweetened beverages as a coping mechanism (20, 22). Given this was a cross-sectional study which could not determine temporality of the associations, we can only speculate about the directions of the relationships.

Our regression models did not show associations between odds of depression and anxiety and fruits, vegetables, legumes, wholegrains, nuts & seeds, milk, red meat, processed meat, fibre, calcium, sodium, PUFA, and trans-fats or other sugar types (fructose, galactose, glucose, lactose, maltose). However, participants with depression and anxiety appeared to have different dietary intakes compared to participants without these conditions. This discrepancy may be a result of insufficient power to detect associations in logistic regression models. This is supported by the associations we did observe between various dietary exposures and continuous depression and anxiety scores (see Supplementary Table 7 in Supplementary Materials). Furthermore, the sample was predominantly a homogeneous sample of university students, therefore the little variance in the dietary exposures might have reduced the possibility of significant findings. Increased awareness of the impact of food on health in this subgroup of educated participants might have also influenced our largely null findings (49).

The current study is not free from limitations. Firstly, due to the cross-sectional nature of this study, we cannot make establish causality, and a possibility of reverse causation cannot be excluded. In fact, as mentioned above, it is well known that depression and anxiety may lead to higher sugar intake (50), possibly due to emotional eating (21), and so they may be more likely to select sweet foods/sugar-sweetened beverages as a coping mechanism (22). Therefore, gender-matched longitudinal designs (considering the gender disproportionality in the current sample) should be adopted in the future to clarify the directionality of these associations. Secondly, participants responding to FFQs are known to be prone to misreporting (51) and might also be affected to recall bias. Other self-reported data, such as height and weight, may likewise introduce inaccuracies. Thirdly, although the DASS subscales predict clinical diagnosis of depression (sensitivity and specificity of 84%) and anxiety (sensitivity of 74% and a specificity of 84% (52)), total score cut-off points on the DASS subscales do not correspond with a clinical diagnosis, but rather indicate the level of high depressive and/or anxiety symptoms self-reported by the individual that may be of clinical relevance. Hence, future studies should consider conducting diagnostic psychiatric interviews to assess depression and anxiety. Fourthly, the sample primarily consisted of university students and there were substantial differences in sample size between male and female participants, which may have influenced the generalisability and applicability of the findings. Fifthly, the absence of physiological data limits our ability to fully support the interpretations and assumptions drawn from the findings. Finally, although the models were adjusted for a variety of major potential confounders, it is possible that the presence of unknown/unmeasured dietary or non-dietary factors could partly explain the reported results, indicating potential residual confounding.

In conclusion, the present study adds to the body of evidence that sugar-sweetened beverage and dietary sugar intakes are associated with increased chance of depression and anxiety. Further large scale longitudinal epidemiological studies assessing these associations are warranted to better establish directionality, temporality, and causality.

## Supporting information

STROBE

## Data Availability

Data available on request from the authors.

## Acknowledgements

This manuscript has been prepared in accordance with the requirements of the Global burden of disease Lifestyle And mental Disorder (GLAD) Taskforce, as part of a global collaborative project to inform the Global Burden of Diseases, Injuries, and Risk Factors Study.

