## Supplementary material for "Sugar intake is associated with increased odds of depression and anxiety: Evidence from a cross-sectional study": STROBE

STROBE Statement—Checklist of items that should be included in reports of ***cross-sectional studies***

|  | Item No | Recommendation |
| --- | --- | --- |
| **Title and abstract** | 1 | (*a*) Indicate the study’s design with a commonly used term in the title or the abstract  **Cross-sectional study as stated in the Title, the Abstract on page 2.** |
|  |  | (*b*) Provide in the abstract an informative and balanced summary of what was done and what was found  **Provided in Abstract on page 2.** |
| Introduction | | |
| Background/rationale | 2 | Explain the scientific background and rationale for the investigation being reported  **Included in the Introduction on pages 3 and 4.** |
| Objectives | 3 | State specific objectives, including any prespecified hypotheses  **Included in the Introduction on page 4.** |
| Methods | | |
| Study design | 4 | Present key elements of study design early in the paper  **Included in the Method on pages 4 and 5.** |
| Setting | 5 | Describe the setting, locations, and relevant dates, including periods of recruitment, exposure, follow-up, and data collection  **Included in the Method on pages 4 and 5.** |
| Participants | 6 | (*a*) Give the eligibility criteria, and the sources and methods of selection of participants  **Included in the Method on pages 4 and 5.** |
| Variables | 7 | Clearly define all outcomes, exposures, predictors, potential confounders, and effect modifiers. Give diagnostic criteria, if applicable  **Included in the Method on pages 4 and 5.** |
| Data sources/ measurement | 8* | For each variable of interest, give sources of data and details of methods of assessment (measurement). Describe comparability of assessment methods if there is more than one group  **Included in the Method on pages 4 and 5.** |
| Bias | 9 | Describe any efforts to address potential sources of bias  **Addressed in the limitations paragraph in the Discussion on page 16.** |
| Study size | 10 | Explain how the study size was arrived at  **Included in the Method on pages 4 and 5.** |
| Quantitative variables | 11 | Explain how quantitative variables were handled in the analyses. If applicable, describe which groupings were chosen and why  **Included in the Method on pages 4 and 5.** |
| Statistical methods | 12 | (*a*) Describe all statistical methods, including those used to control for confounding  **Included in the Method on pages 4 and 5.** |
|  |  | (*b*) Describe any methods used to examine subgroups and interactions  **Included in the Method on pages 4 and 5.** |
|  |  | (*c*) Explain how missing data were addressed  **Included in the Method on pages 4 and 5 and Supplementary Materials.** |
|  |  | (*d*) If applicable, describe analytical methods taking account of sampling strategy **Not applicable.** |
|  |  | (*e*) Describe any sensitivity analyses  **Included in the Method on pages 4 and 5 and Supplementary Materials.** |
| Results | | |
| Participants | 13* | (a) Report numbers of individuals at each stage of study—eg numbers potentially eligible, examined for eligibility, confirmed eligible, included in the study, completing follow-up, and analysed  **Included in the Method on page 4 and in the Results on page 5.** |
|  |  | (b) Give reasons for non-participation at each stage  **Not applicable.** |
|  |  | (c) Consider use of a flow diagram  **Not required.** |
| Descriptive data | 14* | (a) Give characteristics of study participants (eg demographic, clinical, social) and information on exposures and potential confounders  **Included in the Results on pages 5, 6, and 7.** |
|  |  | (b) Indicate number of participants with missing data for each variable of interest  **Included in the Method on pages 4 and 5 and Supplementary Materials.** |
| Outcome data | 15* | Report numbers of outcome events or summary measures  **Included in the Results on pages 5, 6, 7, and 8.** |
| Main results | 16 | (*a*) Give unadjusted estimates and, if applicable, confounder-adjusted estimates and their precision (eg, 95% confidence interval). Make clear which confounders were adjusted for and why they were included  **Included in the Method on page 5 and in the Results on pages 8 to 12.** |
|  |  | (*b*) Report category boundaries when continuous variables were categorized  **Included in the Methods on page 5.** |
|  |  | (*c*) If relevant, consider translating estimates of relative risk into absolute risk for a meaningful time period  **Not relevant.** |
| Other analyses | 17 | Report other analyses done—eg analyses of subgroups and interactions, and sensitivity analyses  **Included in the Results on pages 8 and 9 and Supplementary Materials.** |
| Discussion | | |
| Key results | 18 | Summarise key results with reference to study objectives  **Included in the Discussion on page 13.** |
| Limitations | 19 | Discuss limitations of the study, taking into account sources of potential bias or imprecision. Discuss both direction and magnitude of any potential bias  **Included in the Discussion on page 15.** |
| Interpretation | 20 | Give a cautious overall interpretation of results considering objectives, limitations, multiplicity of analyses, results from similar studies, and other relevant evidence  **Included in the Discussion on pages 13 to 15.** |
| Generalisability | 21 | Discuss the generalisability (external validity) of the study results  **Included in the Method on page 5 and in the Discussion on page 15.** |
| Other information | | |
| Funding | 22 | Give the source of funding and the role of the funders for the present study and, if applicable, for the original study on which the present article is based  **Provided in the Financial Support on the Title page.** |

*Give information separately for exposed and unexposed groups.
