## Supplementary Materials for "Sugar intake is associated with increased odds of depression and anxiety: Evidence from a cross-sectional study"

**Supplementary Note 1: Data cleaning**

From the original sample (n=532), after accounting for missing data; energy intake (n=74) and/or DASS (n=54), a further n=78 participants were excluded if energy intakes (kcal/day) were reported as <500 or >3500kcal per day (Banna et al., 2017). Three participants with energy intakes identified as outliers (>3SD from the mean) were also excluded, resulting in a final sample size of n=377 for analysis.

**Supplementary Table 1: Aligning EPIC output to GLAD exposure variables**

| **GLAD exposure variable** | **EPIC Norfolk output** | **Recalculation/mapped variables/standardisation** |
| --- | --- | --- |
| Veg (g) | Vegetables (g) | Sweetcorn, baked beans and dried lentils, beans, peas removed from Vegetables |
| Legumes (g) | n/a | Intakes of sweetcorn, baked beans and dried lentils, beans, peas |
| Milk (g) | Milk (pints per day) | Milk intake |
| Red meat (g) | Meat & meat products (g) | Intakes of beef, pork, lamb, and goat |
| Processed red meat (g) | Meat & meat products (g) | Intakes of bacon, ham, corned beef, spam, luncheon meats, sausages, savoury pies |
| Sweet drinks (g) | n/a | Intake of fizzy soft drinks and fruit squash or cordial |
| Calcium (g) | calcium (mg) | Milligrams to grams |
| PUFA (% of total energy intake) | PUFA | Grams to % of total energy intake |
| Sodium (g) | sodium (mg) | Milligrams to grams |

**Supplementary Note 2: Estimating Trans-fat intakes**

To address the limitation of trans-fat calculation within the analysis software, FETA, we contacted the EPIC Norfolk team. They provided us with a nutrient database containing trans-fat content specifically for the food items included in the EPIC Norfolk FFQ (Bingham et al., 1997).

Food items from trans-fat nutrient database were pre-screened against the FETA food items ([mrc-epid.cam.ac.uk/wp-content/uploads/2023/03/FFQ_portion_sizes.pdf](https://www.mrc-epid.cam.ac.uk/wp-content/uploads/2023/03/FFQ_portion_sizes.pdf)). This allowed us to identify trans-fat content in the 53 food items. These items were then matched to corresponding entries in the database for further analysis. Frequency categories reported in the FFQ (e.g., 'once a week') were then converted into a 'portion multiplier' (e.g., 0.14 for once a week). This multiplier was then multiplied by the portion size reported in the FFQ to obtain an average daily food weight for each food item. Daily trans-fat intake was then calculated by multiplying the average daily food weight by the nutrient composition per gram and summed across each participant to obtain their average daily trans-fat intake. Supplementary Table 2 includes a list of food items included in trans-fat calculation.

**Supplementary Table 2: Food items from FFQ included in estimation of trans fat.**

| Milk | Potato salad | Low-fat spread |
| --- | --- | --- |
| Beef | Lasagne | Very low-fat spread |
| Burger | Pizza | Chocolate biscuit |
| Pork | Single cream | Plain biscuit |
| Lamb | Double cream | Readymade fruit pies |
| Chicken | Low-fat yogurt | Milk puddings |
| Bacon | Full fat yogurt | Ice cream |
| Ham | Cheese | Chocolates |
| Corned beef | Cottage cheese | Chocolate bars |
| Sausages | Quiche | Crisps |
| Savoury pies | Low calorie salad cream | Vegetable soup |
| Fried fish | Salad cream | Meat soup |
| Roe | French | Peanut butter |
| White bread | Other dressing | Coffee whitener |
| Brown bread | Butter | Coleslaw |
| Wholemeal bread | Hard margarine | Tofu |
| Crackers | Polyunsaturated margarine | Eggs |
| Porridge | Other margarine | Dairy dessert |

**Supplementary Table 3.** Sensitivity analyses of the associations between energy adjusted sugar intake, depression, and anxiety

| **Depression** | | | | | | | | | | | | | |
| --- | --- | --- | --- | --- | --- | --- | --- | --- | --- | --- | --- | --- | --- |
|  | **Model 1*** | | | | **Model 2***** | | | | | **Model 3***** | | | |
|  | **OR** | **p** | **95%CI**  **Lower/Upper** | | **OR** | **P*adj*** | **95%CI**  **Lower/Upper** | | **OR** | | **P*adj*** | **95%CI**  **Lower/Upper** | |
| **Sugars total** | **1.37** | 0.008 | 1.02 | 1.85 | 1.40 | 0.09 | 1.02 | 1.92 | 1.40 | | 0.08 | 1.02 | 1.92 |
|  | **Model 1** | | | | **Model 2***** | | | | | **Model 3***** | | | |
|  | **OR** | **P*adj*** | **95%CI**  **Lower/Upper** | | **OR** | **P*adj*** | **95%CI**  **Lower/Upper** | | **OR** | | **P*adj*** | **95%CI**  **Lower/Upper** | |
| **Fructose** | 0.79 | 0.17 | 0.22 | 2.87 | 0.84 | 0.19 | 0.21 | 3.38 | 0.82 | | 0.19 | 0.20 | 3.31 |
| **Galactose** | 0.63 | 0.04 | 0.37 | 1.05 | 0.69 | 0.11 | 0.42 | 1.16 | 0.69 | | 0.10 | 0.41 | 1.15 |
| **Glucose** | 1.41 | 0.13 | 0.38 | 5.31 | 1.46 | 0.17 | 0.35 | 6.09 | 1.51 | | 0.16 | 0.36 | 6.33 |
| **Lactose** | 1.02 | 0.21 | 0.73 | 1.43 | 1.02 | 0.22 | 0.72 | 1.45 | 1.02 | | 0.22 | 0.71 | 1.44 |
| **Maltose** | 0.99 | 0.25 | 0.54 | 1.80 | 0.98 | 0.23 | 0.51 | 1.86 | 0.98 | | 0.24 | 0.51 | 1.86 |
| **Sucrose** | 1.26 | 0.08 | 0.80 | 1.98 | 1.23 | 0.16 | 0.76 | 1.98 | 1.22 | | 0.15 | 0.76 | 1.97 |
| **Anxiety** | | | | | | | | | | | | | |
|  | **Model 1*** | | | | **Model 2***** | | | | | **Model 3***** | | | |
|  | **OR** | **p** | **95%CI**  **Lower/Upper** | | **OR** | **P*adj*** | **95%CI**  **Lower/Upper** | | **OR** | | **P*adj*** | **95%CI**  **Lower/Upper** | |
| **Sugars total** | **1.35** | <0.001 | 1.03 | 1.78 | 1.36 | 0.09 | 1.01 | 1.83 | 1.36 | | 0.08 | 1.01 | 1.82 |
|  | **Model 1** | | | | **Model 2***** | | | | | **Model 3***** | | | |
|  | **OR** | **P*adj*** | **95%CI**  **Lower/Upper** | | **OR** | **P*adj*** | **95%CI**  **Lower/Upper** | | **OR** | | **P*adj*** | **95%CI**  **Lower/Upper** | |
| **Fructose** | 2.23 | 0.13 | 0.73 | 6.82 | 2.68 | 0.09 | 0.77 | 9.36 | 2.92 | | 0.07 | 0.83 | 10.26 |
| **Galactose** | 0.80 | 0.17 | 0.55 | 1.19 | 0.83 | 0.19 | 0.53 | 1.31 | 0.84 | | 0.19 | 0.53 | 1.31 |
| **Glucose** | 0.42 | 0.08 | 0.13 | 1.37 | 0.37 | 0.11 | 0.10 | 1.40 | 0.34 | | 0.10 | 0.09 | 1.30 |
| **Lactose** | 1.05 | 0.25 | 0.77 | 1.42 | 1.12 | 0.22 | 0.79 | 1.59 | 1.14 | | 0.21 | 0.80 | 1.63 |
| **Maltose** | 1.21 | 0.21 | 0.70 | 2.11 | 1.15 | 0.24 | 0.62 | 2.14 | 1.14 | | 0.24 | 0.61 | 2.12 |
| **Sucrose** | **1.48** | 0.04 | 0.97 | 2.26 | 1.51 | 0.06 | 0.94 | 2.41 | 1.53 | | 0.06 | 0.95 | 2.45 |

* p≤ 0.05, ** p≤ 0.01, p≤ 0.001; *Model 1: unadjusted; Model 2: adjusted for age, sex, income; and Model 3: as in model 2, with body mass index (BMI; kg/m2)*

**Supplementary Table 4.** Sensitivity analyses of the associations between sugar intake, depression, and anxiety in the whole sample (prior to participant exclusion)

| **Depression** | | | | | | | | | | | | | |
| --- | --- | --- | --- | --- | --- | --- | --- | --- | --- | --- | --- | --- | --- |
|  | **Model 1***** | | | | **Model 2***** | | | | | **Model 3***** | | | |
|  | **OR** | **p** | **95%CI**  **Lower/Upper** | | **OR** | **P*adj*** | **95%CI**  **Lower/Upper** | | **OR** | | **P*adj*** | **95%CI**  **Lower/Upper** | |
| **Sugars total** | **1.00** | <0.001 | 1.00 | 1.00 | **1.00** | 0.02 | 1.00 | 1.00 | **1.00** | | 0.02 | 1.00 | 1.00 |
|  | **Model 1***** | | | | **Model 2***** | | | | | **Model 3***** | | | |
|  | **OR** | **P*adj*** | **95%CI**  **Lower/Upper** | | **OR** | **P*adj*** | **95%CI**  **Lower/Upper** | | **OR** | | **P*adj*** | **95%CI**  **Lower/Upper** | |
| **Fructose** | 0.98 | 0.17 | 0.91 | 1.05 | 1.00 | 0.21 | 0.92 | 1.08 | 1.00 | | 0.18 | 0.92 | 1.08 |
| **Galactose** | **0.79** | 0.04 | 0.59 | 1.08 | 0.73 | 0.06 | 0.52 | 1.03 | 0.73 | | 0.06 | 0.52 | 1.03 |
| **Glucose** | 1.03 | 0.13 | 0.95 | 1.11 | 1.01 | 0.16 | 0.92 | 1.10 | 1.01 | | 0.17 | 0.92 | 1.10 |
| **Lactose** | 1.01 | 0.21 | 0.97 | 1.05 | 1.02 | 0.15 | 0.98 | 1.05 | 1.01 | | 0.14 | 0.98 | 1.05 |
| **Maltose** | 1.11 | 0.08 | 0.96 | 1.28 | 1.07 | 0.13 | 0.92 | 1.24 | 1.07 | | 0.12 | 0.92 | 1.25 |
| **Sucrose** | 1.00 | 0.25 | 0.99 | 1.01 | 1.00 | 0.12 | 0.99 | 1.01 | 1.00 | | 0.11 | 0.99 | 1.01 |
| **Anxiety** | | | | | | | | | | | | | |
|  | **Model 1***** | | | | **Model 2***** | | | | | **Model 3***** | | | |
|  | **OR** | **p** | **95%CI**  **Lower/Upper** | | **OR** | **P*adj*** | **95%CI**  **Lower/Upper** | | **OR** | | **P*adj*** | **95%CI**  **Lower/Upper** | |
| **Sugars total** | **1.01** | <0.001 | 1.01 | 1.01 | **1.01** | 0.02 | 1.01 | 1.01 | **1.01** | | 0.02 | 1.01 | 1.01 |
|  | **Model 1***** | | | | **Model 2***** | | | | | **Model 3***** | | | |
|  | **OR** | **P*adj*** | **95%CI**  **Lower/Upper** | | **OR** | **P*adj*** | **95%CI**  **Lower/Upper** | | **OR** | | **P*adj*** | **95%CI**  **Lower/Upper** | |
| **Fructose** | 1.06 | 0.17 | 0.98 | 1.15 | 1.05 | 0.12 | 0.97 | 1.15 | 1.07 | | 0.11 | 0.98 | 1.16 |
| **Galactose** | 1.01 | 0.25 | 0.74 | 1.39 | 1.06 | 0.24 | 0.75 | 1.51 | 1.06 | | 0.24 | 0.75 | 1.50 |
| **Glucose** | 0.94 | 0.13 | 0.85 | 1.02 | 0.94 | 0.13 | 0.85 | 1.04 | 0.93 | | 0.14 | 0.84 | 1.03 |
| **Lactose** | 1.01 | 0.21 | 0.97 | 1.05 | 1.02 | 0.18 | 0.97 | 1.06 | 1.02 | | 0.18 | 0.98 | 1.06 |
| **Maltose** | 1.17 | 0.08 | 0.99 | 1.40 | 1.10 | 0.15 | 0.91 | 1.33 | 1.10 | | 0.15 | 0.91 | 1.33 |
| **Sucrose** | **1.01** | 0.04 | 1.00 | 1.03 | 1.02 | 0.06 | 1.00 | 1.03 | 1.02 | | 0.06 | 1.00 | 1.03 |

* p≤ 0.05, ** p≤ 0.01, p≤ 0.001; *Model 1: unadjusted; Model 2: adjusted for age, sex, income; and Model 3: as in model 2, with body mass index (BMI; kg/m2)*

**Supplementary Table 5.** Sensitivity analyses of the associations between energy adjusted dietary variables, depression, and anxiety

| **Depression** | | | | | | | | | | | | | | |
| --- | --- | --- | --- | --- | --- | --- | --- | --- | --- | --- | --- | --- | --- | --- |
|  | **Model 1** | | | | | **Model 2**** | | | | | **Model 3**** | | | |
|  | **OR** | **P*adj*** | **95%CI**  **Lower/Upper** | | **OR** | | **P*adj*** | **95%CI**  **Lower/Upper** | | **OR** | | **P*adj*** | **95%CI**  **Lower/Upper** | |
| **Fruits** | 1.13 | 0.14 | 0.70 | 1.81 | 0.95 | | 0.21 | 0.59 | 1.53 | 0.95 | | 0.19 | 0.59 | 1.53 |
| **Vegetables** | 1.06 | 0.25 | 0.52 | 2.15 | 0.96 | | 0.22 | 0.44 | 2.09 | 0.96 | | 0.21 | 0.44 | 2.09 |
| **Legumes** | 0.67 | 0.04 | 0.41 | 1.12 | 0.68 | | 0.07 | 0.42 | 1.12 | 0.68 | | 0.07 | 0.42 | 1.12 |
| **Wholegrains** | 0.92 | 0.16 | 0.62 | 1.38 | 0.83 | | 0.11 | 0.55 | 1.25 | 0.83 | | 0.11 | 0.55 | 1.25 |
| **Nuts & Seeds** | 1.18 | 0.11 | 0.78 | 1.80 | 1.39 | | 0.09 | 0.90 | 2.14 | 1.39 | | 0.08 | 0.90 | 2.15 |
| **Milk** | 1.07 | 0.20 | 0.63 | 1.80 | 1.14 | | 0.19 | 0.65 | 1.98 | 1.14 | | 0.17 | 0.65 | 1.99 |
| **Red meat** | 1.21 | 0.05 | 0.84 | 1.74 | 1.25 | | 0.10 | 0.85 | 1.83 | 1.25 | | 0.10 | 0.85 | 1.84 |
| **Processed meat** | 1.06 | 0.18 | 0.74 | 1.50 | 1.14 | | 0.14 | 0.78 | 1.67 | 1.14 | | 0.14 | 0.78 | 1.67 |
| **Sugar-sweetened beverages** | 1.43 | 0.01 | 1.06 | 1.94 | **1.34** | | 0.04 | 0.98 | 1.83 | **1.34** | | 0.04 | 0.98 | 1.83 |
| **Fibre** | 0.92 | 0.23 | 0.38 | 2.21 | 1.04 | | 0.24 | 0.41 | 2.64 | 1.04 | | 0.24 | 0.41 | 2.64 |
| **Calcium** | 0.83 | 0.09 | 0.52 | 1.31 | 0.89 | | 0.18 | 0.54 | 1.48 | 0.89 | | 0.18 | 0.54 | 1.48 |
| **Sodium** | 0.96 | 0.07 | 0.62 | 1.47 | 1.00 | | 0.13 | 0.64 | 1.59 | 1.00 | | 0.13 | 0.64 | 1.59 |
| **PUFA** | 1.18 | 0.21 | 0.76 | 1.84 | 1.13 | | 0.25 | 0.70 | 1.81 | 1.13 | | 0.25 | 0.70 | 1.82 |
| **Trans-fats** | 1.19 | 0.13 | 0.80 | 1.75 | 1.18 | | 0.16 | 0.78 | 1.77 | 1.18 | | 0.15 | 0.78 | 1.78 |
| **Anxiety** | | | | | | | | | | | | | | |
|  | **Model 1** | | | | | **Model 2***** | | | | | **Model 3***** | | | |
|  | **OR** | **P*adj*** | **95%CI**  **Lower/Upper** | | **OR** | | **P*adj*** | **95%CI**  **Lower/Upper** | | **OR** | | **P*adj*** | **95%CI**  **Lower/Upper** | |
| **Fruits** | 1.12 | 0.11 | 0.74 | 1.71 | 1.01 | | 0.25 | 0.67 | 1.54 | 1.01 | | 0.24 | 0.67 | 1.54 |
| **Vegetables** | 1.14 | 0.14 | 0.62 | 2.10 | 1.11 | | 0.19 | 0.56 | 2.21 | 1.11 | | 0.19 | 0.56 | 2.19 |
| **Legumes** | 0.84 | 0.09 | 0.56 | 1.24 | 0.85 | | 0.09 | 0.58 | 1.26 | 0.84 | | 0.07 | 0.56 | 1.24 |
| **Wholegrains** | 0.97 | 0.20 | 0.68 | 1.37 | 0.93 | | 0.18 | 0.65 | 1.33 | 0.94 | | 0.17 | 0.65 | 1.34 |
| **Nuts & Seeds** | 0.90 | 0.13 | 0.59 | 1.35 | 1.10 | | 0.16 | 0.71 | 1.69 | 1.11 | | 0.13 | 0.72 | 1.72 |
| **Milk** | 1.07 | 0.18 | 0.67 | 1.71 | 1.20 | | 0.10 | 0.73 | 1.97 | 1.24 | | 0.10 | 0.75 | 2.05 |
| **Red meat** | 0.84 | 0.07 | 0.59 | 1.20 | 0.90 | | 0.12 | 0.62 | 1.31 | 0.93 | | 0.18 | 0.64 | 1.36 |
| **Processed meat** | 1.00 | 0.25 | 0.71 | 1.40 | 1.05 | | 0.21 | 0.72 | 1.54 | 1.04 | | 0.21 | 0.71 | 1.52 |
| **Sugar-sweetened beverages** | 1.2 | 0.02 | 0.91 | 1.62 | 1.15 | | 0.06 | 0.85 | 1.55 | 1.14 | | 0.08 | 0.84 | 1.55 |
| **Fibre** | 0.86 | 0.16 | 0.41 | 1.81 | 0.92 | | 0.22 | 0.41 | 2.05 | 0.92 | | 0.22 | 0.41 | 2.08 |
| **Calcium** | 0.82 | 0.05 | 0.54 | 1.22 | 0.89 | | 0.13 | 0.56 | 1.41 | 0.91 | | 0.14 | 0.58 | 1.45 |
| **Sodium** | 1.00 | 0.21 | 0.69 | 1.44 | 1.10 | | 0.24 | 0.74 | 1.63 | 1.08 | | 0.25 | 0.73 | 1.60 |
| **PUFA** | 1.24 | 0.23 | 0.83 | 1.85 | 1.20 | | 0.15 | 0.78 | 1.82 | 1.15 | | 0.15 | 0.75 | 1.76 |
| **Trans-fats** | 0.99 | 0.04 | 0.69 | 1.42 | 0.98 | | 0.07 | 0.67 | 1.43 | 1.01 | | 0.11 | 0.69 | 1.47 |

* p≤ 0.05, ** p≤ 0.01, p≤ 0.001; *Model 1: unadjusted; Model 2: adjusted for age, sex, income; and Model 3: as in model 2, with body mass index (BMI; kg/m2)*

**Supplementary Table 6.** Sensitivity analyses of the associations between dietary variables, depression, and anxiety in the whole sample (prior to participant exclusion)

| **Depression** | | | | | | | | | | | | | | |
| --- | --- | --- | --- | --- | --- | --- | --- | --- | --- | --- | --- | --- | --- | --- |
|  | **Model 1***** | | | | | **Model 2***** | | | | | **Model 3***** | | | |
|  | **OR** | **P*adj*** | **95%CI**  **Lower/Upper** | | **OR** | | **P*adj*** | **95%CI**  **Lower/Upper** | | **OR** | | **P*adj*** | **95%CI**  **Lower/Upper** | |
| **Fruits** | 1.00 | 0.18 | 1.00 | 1.00 | 1.00 | | 0.13 | 1.00 | 1.00 | 1.00 | | 0.13 | 1.00 | 1.00 |
| **Vegetables** | 1.00 | 0.14 | 1.00 | 1.00 | 1.00 | | 0.10 | 1.00 | 1.00 | 1.00 | | 0.10 | 1.00 | 1.00 |
| **Legumes** | 1.00 | 0.11 | 0.99 | 1.01 | 1.00 | | 0.16 | 0.99 | 1.01 | 1.00 | | 0.15 | 0.99 | 1.01 |
| **Wholegrains** | 1.00 | 0.13 | 1.00 | 1.00 | 1.00 | | 0.06 | 1.00 | 1.00 | 1.00 | | 0.06 | 1.00 | 1.00 |
| **Nuts & Seeds** | 1.00 | 0.23 | 0.98 | 1.02 | 1.00 | | 0.24 | 0.98 | 1.02 | 1.00 | | 0.22 | 0.98 | 1.02 |
| **Milk** | **1.00** | 0.04 | 1.00 | 1.00 | 1.00 | | 0.07 | 1.00 | 1.00 | 1.00 | | 0.07 | 1.00 | 1.00 |
| **Red meat** | **1.00** | 0.05 | 1.00 | 1.01 | 1.00 | | 0.18 | 1.00 | 1.00 | 1.00 | | 0.17 | 1.00 | 1.00 |
| **Processed meat** | 1.00 | 0.07 | 0.99 | 1.01 | 1.00 | | 0.12 | 0.99 | 1.01 | 1.00 | | 0.11 | 0.99 | 1.01 |
| **Sugar-sweetened beverages** | **1.00** | 0.02 | 1.00 | 1.01 | **1.00** | | 0.04 | 1.00 | 1.00 | **1.00** | | 0.04 | 1.00 | 1.00 |
| **Fibre** | 1.01 | 0.16 | 0.94 | 1.09 | 1.03 | | 0.09 | 0.95 | 1.12 | 1.03 | | 0.08 | 0.95 | 1.12 |
| **Calcium** | 1.08 | 0.21 | 0.38 | 3.10 | 1.45 | | 0.15 | 0.50 | 4.23 | 1.45 | | 0.14 | 0.50 | 4.23 |
| **Sodium** | 0.98 | 0.09 | 0.83 | 1.16 | 1.01 | | 0.22 | 0.84 | 1.21 | 1.01 | | 0.25 | 0.84 | 1.21 |
| **PUFA** | 1.00 | 0.20 | 1.00 | 1.00 | 1.00 | | 0.22 | 1.00 | 1.00 | 1.00 | | 0.21 | 1.00 | 1.00 |
| **Trans-fats** | 1.00 | 0.25 | 0.19 | 5.33 | 0.81 | | 0.21 | 0.14 | 4.61 | 0.81 | | 0.19 | 0.14 | 4.60 |
| **Anxiety** | | | | | | | | | | | | | | |
|  | **Model 1***** | | | | | **Model 2***** | | | | | **Model 3***** | | | |
|  | **OR** | **P*adj*** | **95%CI**  **Lower/Upper** | | **OR** | | **P*adj*** | **95%CI**  **Lower/Upper** | | **OR** | | **P*adj*** | **95%CI**  **Lower/Upper** | |
| **Fruits** | 1.00 | 0.13 | 1.00 | 1.00 | 1.00 | | 0.13 | 1.00 | 1.00 | 1.00 | | 0.13 | 1.00 | 1.00 |
| **Vegetables** | 1.00 | 0.07 | 1.00 | 1.00 | 1.00 | | 0.06 | 1.00 | 1.00 | 1.00 | | 0.07 | 1.00 | 1.00 |
| **Legumes** | 1.01 | 0.09 | 1.00 | 1.02 | 1.01 | | 0.12 | 1.00 | 1.02 | 1.00 | | 0.14 | 0.99 | 1.02 |
| **Wholegrains** | 1.00 | 0.25 | 1.00 | 1.00 | 1.00 | | 0.21 | 1.00 | 1.00 | 1.00 | | 0.21 | 1.00 | 1.00 |
| **Nuts & Seeds** | 0.99 | 0.16 | 0.97 | 1.02 | 1.00 | | 0.24 | 0.98 | 1.03 | 1.01 | | 0.24 | 0.98 | 1.03 |
| **Milk** | 1.00 | 0.20 | 1.00 | 1.00 | 1.00 | | 0.18 | 1.00 | 1.00 | 1.00 | | 0.18 | 1.00 | 1.00 |
| **Red meat** | **1.00** | 0.05 | 1.00 | 1.01 | 1.00 | | 0.10 | 1.00 | 1.01 | 1.00 | | 0.08 | 1.00 | 1.01 |
| **Processed meat** | 1.00 | 0.11 | 0.99 | 1.01 | 1.00 | | 0.25 | 0.99 | 1.01 | 1.00 | | 0.25 | 0.99 | 1.01 |
| **Sugar-sweetened beverages** | **1.00** | 0.02 | 1.00 | 1.01 | 1.00 | | 0.07 | 1.00 | 1.01 | 1.00 | | 0.01 | 1.00 | 1.01 |
| **Fibre** | 1.03 | 0.14 | 0.95 | 1.12 | 1.04 | | 0.16 | 0.95 | 1.14 | 1.04 | | 0.15 | 0.95 | 1.15 |
| **Calcium** | **2.92** | 0.04 | 0.84 | 10.15 | **4.72** | | 0.04 | 1.20 | 18.58 | **4.84** | | 0.04 | 1.23 | 18.95 |
| **Sodium** | 1.01 | 0.18 | 0.86 | 1.20 | 1.08 | | 0.22 | 0.91 | 1.29 | 1.08 | | 0.22 | 0.91 | 1.29 |
| **PUFA** | 1.00 | 0.21 | 1.00 | 1.00 | 1.00 | | 0.15 | 1.00 | 1.00 | 1.00 | | 0.17 | 1.00 | 1.00 |
| **Trans-fats** | 0.91 | 0.23 | 0.16 | 5.21 | 0.60 | | 0.19 | 0.10 | 3.56 | 0.56 | | 0.19 | 0.10 | 3.21 |

* p≤ 0.05, ** p≤ 0.01, p≤ 0.001; *Model 1: unadjusted; Model 2: adjusted for age, sex, income; and Model 3: as in model 2, with body mass index (BMI; kg/m2)*

**Supplementary Table 7.** Associations between continuous depression and anxiety scores and dietary exposures

|  | **Depression** | | | | | | **Anxiety** | | | |
| --- | --- | --- | --- | --- | --- | --- | --- | --- | --- | --- |
|  | **Pearson**  **Correlation** | **P*adj*** | **95%CI**  **Lower/Upper** | | | **Pearson**  **Correlation** | | **P*adj*** | **95%CI**  **Lower/Upper** | |
| **Fruits** | -.006 | 0.23 | -0.11 | 0.10 | .014 | | | 0.15 | -0.09 | 0.11 |
| **Vegetables** | -.069 | 0.12 | -0.17 | 0.03 | -.045 | | | 0.23 | -0.15 | 0.06 |
| **Legumes** | -.078 | 0.10 | -0.18 | 0.02 | -.042 | | | 0.17 | -0.14 | 0.06 |
| **Wholegrains** | .050 | 0.13 | -0.05 | 0.15 | .105 | | | 0.07 | 0.00 | 0.20 |
| **Nuts & Seeds** | -.015 | 0.20 | -0.12 | 0.09 | -.097 | | | 0.08 | -0.20 | 0.00 |
| **Milk** | -.020 | 0.17 | -0.12 | 0.08 | .017 | | | 0.22 | -0.08 | 0.12 |
| **Red meat** | **.150** | 0.03 | 0.05 | 0.25 | **.111** | | | 0.05 | 0.01 | 0.21 |
| **Processed meat** | .096 | 0.07 | -0.01 | 0.20 | .031 | | | 0.18 | -0.07 | 0.13 |
| **Sugar-sweetened beverages** | **.189** | 0.02 | 0.09 | 0.29 | **.165** | | | 0.02 | 0.07 | 0.26 |
| **Fibre** | -.013 | 0.22 | -0.11 | 0.09 | .021 | | | 0.20 | -0.08 | 0.12 |
| **Calcium** | .017 | 0.18 | -0.08 | 0.12 | .071 | | | 0.12 | -0.03 | 0.17 |
| **Sodium** | .083 | 0.08 | -0.02 | 0.18 | **.135** | | | 0.03 | 0.03 | 0.23 |
| **PUFA** | **-.103** | 0.05 | -0.20 | 0.00 | -.082 | | | 0.10 | -0.18 | 0.02 |
| **Trans fats** | .046 | 0.15 | -0.08 | 0.13 | .069 | | | 0.13 | -0.04 | 0.16 |
| **Sugars total** | **0.21** | 0.03 | 0.79 | 0.85 | **0.26** | | | 0.03 | 0.17 | 0.34 |
| ***Fructose*** | **0.16** | 0.03 | 0.12 | 0.29 | **0.22** | | | 0.03 | 0.13 | 0.31 |
| ***Galactose*** | 0.11 | 0.06 | 0.07 | 0.25 | **0.16** | | | 0.03 | 0.07 | 0.25 |
| ***Glucose*** | **0.18** | 0.03 | 0.02 | 0.20 | **0.23** | | | 0.03 | 0.14 | 0.32 |
| ***Lactose*** | **0.19** | 0.03 | 0.09 | 0.27 | **0.23** | | | 0.03 | 0.15 | 0.32 |
| ***Maltose*** | **0.25** | 0.03 | 0.10 | 0.27 | **0.30** | | | 0.03 | 0.22 | 0.38 |
| ***Sucrose*** | **0.21** | 0.03 | 0.17 | 0.34 | **0.26** | | | 0.03 | 0.17 | 0.34 |
